# Biological Factors Influencing Individual Responses to Propylene Oxide: A Systematic Review of Exogenous Exposure, Endogenous Production and Detoxification

**DOI:** 10.1101/2024.02.15.24302622

**Authors:** Irene S. Gabashvili

**Affiliations:** Aurametrix, MEBO Research

**Keywords:** Propylene Oxide, Propylene, Human Exhaled Breath, Systematic Review, Environmental Health, AI in Literature Review, Analytical Methodologies, Metabolism

## Abstract

**Objective:** This systematic review aims to synthesize current knowledge on Propylene Oxide (PO) in human exhaled breath, examining its presence across various biological matrices and exploring methodologies for its analysis. It seeks to elucidate the sources of PO in the human body and understand individual variability in detoxification processes.

**Methods:** A comprehensive literature search was conducted across 12 databases and specialized repositories, spanning over 10,000 publications without language restrictions until May 16, 2024. Seventeen AI tools were employed to enhance study identification and analysis, focusing on both direct mentions and indirect evidence of PO behavior and detection in the human body. Assessment tools for risk of bias included SYRCLE’s tool for animal studies, the Newcastle-Ottawa Scale for cohort and case-control studies, and the ROBINS-I tool for non-randomized studies. The selection process yielded 89 studies, encompassing a range of research types and species, supplemented by reviews, monographs, and editorials to provide a comprehensive overview.

**Results:** The search revealed limited direct evidence on PO concentrations in exhaled breath, with only one reference providing concrete data (0.083 ppb to 0.3 ppb following exposures to 10-25 ppm of propylene). A study of ours, published separately, indicated significantly higher PO concentrations (hundreds of ppb or even ppm) in individuals with environmental sensitivities. Numerous references offered indirect insights into PO’s persistence and detection in blood and urine. The review highlights the enzymes involved in PO metabolism, the evolution of analytical methodologies, and the challenges and potential of AI tools in systematic reviews.

**Conclusions:** The scarcity of direct evidence on PO in exhaled breath underscores a significant gap in the literature and existing databases. Directly and indirectly relevant sources indicate variability in environmental compound concentrations in exhaled breath, influenced by genetics, health status, metabolism, and the microbiome. The review emphasizes the difficulties in synthesizing data on PO effects due to heterogeneous inputs and complex exposure scenarios. It underscores the need for advanced AI capabilities in literature reviews to capture nuanced, indirect evidence more effectively and calls for targeted research and technological innovation in environmental health sciences. Enhancing AI tools to navigate scientific literature with greater efficacy can leverage PRISMA guidelines and diverse data sources to minimize bias and enhance reliability. This approach will aid in addressing the complex interplay of biological and environmental factors in PO metabolism and toxicity, ultimately improving risk assessments and intervention strategies.

## Introduction

### Rationale

In our daily lives, we encounter a myriad of chemical substances, from the products we use to the environments we inhabit. Whether it’s in industrial settings where chemicals like propylene oxide are utilized, or in the air we breathe in neighborhoods adjacent to chemical plants, the presence of these substances is pervasive. Even activities like personal care, driving, cleaning, gardening, painting, or exposure to secondary smoke introduce us to a plethora of chemicals.

Propylene oxide, though less notorious than some chemicals, is omnipresent due to its extensive industrial and consumer applications, raising concerns about potential health risks and individual variations in exposure levels. This underscores the need for comprehensive investigations into factors influencing these variations, including genetic makeup, overall health status, and environmental interactions. Despite existing research focusing on exposure biomarkers, gaps persist, highlighting the importance of understanding the less-studied PO’s impacts, as revealed in preliminary breath metabolome studies. Consequently, our study aims to illuminate PO levels in human breath through a systematic review, focusing on uptake, metabolism, and elimination, with an emphasis on individual responses and environmental factors. Ultimately, our goal is to offer insights into individual metabolic variations, alongside strategies for exposure reduction and health risk mitigation.

### Objectives

Our study aims to provide an overview of existing research on PO levels in exhaled breath across various health states, examining factors that could potentially influence these levels, such as environmental exposure and metabolism, alongside methods used for quantification. Additionally, we will explore the role of genetic polymorphisms in PO metabolism. Through synthesizing this information, we aim to offer a comprehensive understanding of PO exposure based on data obtained through exhaled breath analysis and identify avenues for future investigation.

By conducting a systematic review of literature on PO levels in human breath, our research addresses a critical knowledge gap, considering its detectable presence and implications for health risk assessments. Emphasizing uptake, metabolism, and elimination, we will explore individual responses to PO exposure, including genetic differences, overall health, and environmental factors.

Our research goes beyond tracing chemical paths. The goal is to decipher diverse human responses shaped by genetics, microbiome, health, and environment, contributing to a safer, better-understood world.

## Methods

### Study Design

Systematic Review follows the Preferred Reporting Items for Systematic Reviews and MetaAnalysis (PRISMA) guidelines. The protocol is registered in PROSPERO [15], an international prospective register of systematic reviews at the National Institute for Health Research and the Center for Reviews and Dissemination (CRD) at the University of York (CRD42023397159).

The study was designed as an open-source collaborative project via Open Source Science Framework. Registered Identifier: DOI 10.17605/OSF.IO/W7682 The literature search spanned multiple databases, including Medrxiv, EuropePMC and PubMed.

### Research Questions

The following research questions have been formulated to guide this review:

- RQ1: What range of propylene oxide concentrations has been documented in human exhaled breath across different studies?
- RQ2: How do health conditions affect the levels of propylene oxide detected in exhaled breath? (RQ2* How different concentrations of PO, its precursors and derivatives are associated with various health conditions)
- RQ3: Which biological variables are known to modulate individual susceptibility to propylene oxide and other environmental epoxides?
- RQ4: What factors are proposed to influence Propylene Oxide levels in the human body, and what methods are used for quantification?
- RQ5: What genetic polymorphisms could potentially affect propylene oxide metabolism?
- RQ6: How does the human microbiome interact with propylene oxide metabolism, potentially influencing its concentration in exhaled breath?

### Literature Search Strategy

A comprehensive search strategy was designed to scour a wide range of databases, making use of specific search terms to ensure a broad coverage of relevant literature Search strategies are described in the [protocol](https://www.crd.york.ac.uk/prospero/display_record.php?ID=CRD42023397159) registered in the Prospero database.

Starting with the MEDLINE database accessed via PubChem, searches were conducted using the PubChem Compound IDs (CIDs) for propylene oxide and its synonyms (CID 6378) and related compounds, with a specific interest in their presence in breath tests or exhaled breath, in metabolomic and breathomic studies.

The search then moved to PubMed directly, using a combination of MeSH (Medical Subject Headings) terms and key phrases. The query combined terms related to breath tests and the metabolome with propylene oxide and its specific synonyms or related MeSH terms.

Propylene oxide, also known as methyloxirane, has several chemical names including Epoxypropane, Propylene epoxide, 1,2-Propylene oxide, Methyl oxirane, 1,2-Epoxypropane, Propene oxide and Methyl ethylene oxide

(“breath tests“[MeSH Terms] OR “exhaled breath” OR (metabolome[MeSH Terms] AND breath)) AND (“propylene oxide” OR “Epoxy Compounds“[MeSH Terms] OR “2-hydroxypropylmercapturic acid” OR propylene)

For Google Scholar, a similar approach was used, replacing MeSH terms with simple keywords. Both ‘breathomics’ and ‘metabolomics’ were included in the query to ensure comprehensive coverage of the field. Specific synonyms for propylene oxide were also included in the query.

“How is propylene oxide metabolized in the living organism?” “How is propylene metabolized in the living organism?”

Next, the search was expanded to Semantic Scholar with a simpler, more direct query: ‘propylene oxide in exhaled breath’. This broad search term was designed to return a range of results relating to the presence of propylene oxide in exhaled breath.

For Dimensions.AI, we applied the following queries: ‘“propylene oxide” AND “exhaled breath“’ in full data; “propylene oxide” AND metabolism’ in title and abstract and removed duplicates.

Query for EuropePMC was Propylene Oxide ((exhaled breath OR metabolism) NOT (electron microscopy OR manufacturing))

Chemrxiv was searched for “propylene oxide” AND “exhaled breath“

We also used seventeen AI tools including Perplexity.AI, Bing, Bard (now Gemini), Claude and scholarly plugins of Chat GPT 4.0 (Science, Scholarly, LitMaps, etc). Later we reran all searches with custom versions of GPTs (rolling out after November 2023) created for literature searches (Such as [Consensus app GPT](https://consensus.app/), PubmedBuddy, SciSpace) and standalone apps like Scite.ai.

Examples of questions asked were:

“What concentrations of propylene oxide were found in exhaled breath?”

”What scholarly papers mention propylene oxide in exhaled breath?”

“What scholarly papers explain how propylene oxide is metabolized in the living organism and how it is created from compounds such as Propylene?”

Finally, the search strategy included one of the most useful sources Elicit.org, a platform that utilizes language models to aid in literature searches. A direct search for ‘propylene oxide in exhaled breath’ was conducted.’.

Our multi-pronged search strategy incorporates diverse databases to ensure a robust and comprehensive literature review. This includes leveraging resources such as:

The [Comptox database](https://comptox.epa.gov/dashboard/chemical-lists/VOLATILOME) which hosts 1,178 chemicals identified in breath [38].

The Human Breathomics Database (HBDB) is described as the most extensive repository of volatile organic compounds (VOCs) found in human exhaled breath [23]. With data sourced from 2766 publications, the HBDB catalogs 1143 compounds and their connections to 60 diseases, offering invaluable insights into breath-based biomarkers.

The Human Metabolome Database, a free resource providing detailed information on small molecule metabolites, drugs, toxins, and food components. With links to other databases and comprehensive clinical and molecular biology data, the Human Metabolome Database enriches our understanding of metabolic pathways and biomolecular interactions.

We also searched for the most relevant author identified:

“propylene oxide“[nm] AND Filser[author] (15 papers on Pubmed)

Final search was conducted on May 15, 2024, but there was very little differences compared to February 1, 2024, November 1, 2023 and July 25, 2023, the dates of previous comprehensive searches.

### Inclusion and Exclusion Criteria

For this review, we considered all studies reporting on the concentrations of Propylene Oxide in exhaled air and/or investigating factors that may potentially influence these concentrations. We deliberately cast a wide net in our search strategy to incorporate various study designs. The different methodologies we included ranged from descriptive, qualitative, analytical, and experimental studies to quasi-experimental and mixed-methods designs. We anticipated that the most prevalent design would be cross-sectional studies, which typically compare populations with diseases, those at risk, or those exposed to Propylene Oxide to healthy/unexposed control groups.

We excluded any studies that discussed Propylene Oxide in a non-relevant context. For instance, studies that mention Propylene Oxide as a compound used in the preparation of biological samples for electron microscopy, or for the removal of residual ethanol used previously for dehydration, fell outside our scope of interest. We also dismissed studies that did not contribute original information or interpretation. These exclusions ensured a focused review and a meaningful aggregation of information pertinent to the specific topic of interest.

### Study Selection

The initial screening involved evaluating titles and abstracts, followed by a comprehensive review of potentially relevant studies in full text. To ensure relevance and avoid duplication, the titles were carefully verified and any identical titles across different versions of the same article were identified and eliminated, particularly preprint versions. In cases of repeat publication, priority was given to the most recent and comprehensive data. As mentioned previously, Propylene Oxide is used as a dehydrating agent and a transition solvent in the preparation of biological samples for electron microscopy and is one of the most used biocompatible polymers in flexible electronics (Bilbao et al. 2023). Papers mentioning PO in this and other context not relevant to this study were filtered out.

In addition to manual review, the titles were systematically ranked based on the relevance of their corresponding abstracts to the study’s topics, such as “metabolism of propylene oxide.” This ranking was calculated using TF-IDF (term frequency–inverse document frequency) and cosine similarity measures, which gauge the importance of words within documents relative to a larger corpus. Furthermore, text-based clustering methods, including k-means clustering on TF-IDF vectors, were employed to group papers with similar content. TF-IDF, a widely utilized term-weighting scheme, helps prioritize words based on their frequency within documents while accounting for their prevalence across the entire corpus. This approach, commonly employed in information retrieval, text mining, and user modeling, enhances the accuracy and efficiency of study selection processes. According to recent studies, over 80% of text-based recommender systems in digital libraries rely on TF-IDF for information retrieval tasks.

### Data Extraction & Synthesis

In the process of data extraction and synthesis for Propylene Oxide studies, relevant concentrations of propylene oxide or associated chemicals, sources and modes of exposure, settings of experiments or observations, and detailed information about subjects were gathered alongside pertinent health effects or other biological variables. Since the studies were not sufficiently homogeneous, the Higgins index (I²) could not be calculated to assess heterogeneity across most studies. Therefore, a systematic narrative synthesis was conducted to analyze the data comprehensively.

Additionally, VOSViewer was utilized to conduct quantitative analysis and generate visualizations for the creation of a knowledge map. This facilitated the exploration of relationships between variables and provided insights into the patterns and trends present within the data.

## Results

### Historical Overview

Timeline visualization of MESH terms and other keywords associated with papers mentioning “Propylene Oxide” in their abstract or title - sourced from databases such as EuropePMC, PubMed, Dimensions. AI, and others offers a dynamic portrait of the evolution of propylene oxide research.

Initial investigations into propylene oxide predominantly utilized rat models to study genetic mutations and environmental hazards during pregnancy. These studies often employed atmospheric exposure chambers, with a particular focus on respiratory system impacts. Key study parameters included aspects like stereoisomerism, effects on acetaminophen metabolism, food contamination risks, and the use of F344 rats as a model organism. Analytical methods such as UV and FTIR spectrometry were frequently employed. The cytochrome 450 enzyme system was a recurring topic, alongside considerations of skin and surface interactions with propylene oxide.

Initial studies focused on atmospheric exposure and respiratory impacts, utilizing techniques like UV and FTIR spectrometry. After a peak in research activity in 2005 (106 publications in 2005 on Pubmed, while only 80 in 2006), a shift towards molecular analysis ensued, introducing advanced tools like magnetic resonance spectroscopy.

Environmental concerns gained prominence post-2009, extending beyond occupational exposure and food contamination. Industrial production of epoxides, linked to substantial greenhouse gas emissions, prompted exploration of green synthesis, using bacteria in the production of propylene oxide and other mitigation strategies. New research frontiers included DNA damage, biomarkers and brain studies. The integration of quantum theory and algorithms has become more prevalent. 2018 saw the rise of publications about using AI in wearable electronic devices [14], although Propylene Oxide was mostly mentioned in the context of poly(ethylene oxide)-poly(propylene oxide)-poly(ethylene oxide) (PEO-PPO-PEO), promising materials for use in wearables. In the past three years, there has been a notable emergence of studies focusing on the impact of Propylene Oxide within the human body in general population, reflecting the growing recognition of its widespread presence. The use of artificial intelligence is becoming increasingly prevalent in processing experimental data and optimizing various aspects of chemical reactions, formulations, and engineering processes, including exploration and production.

**Figure 1.**
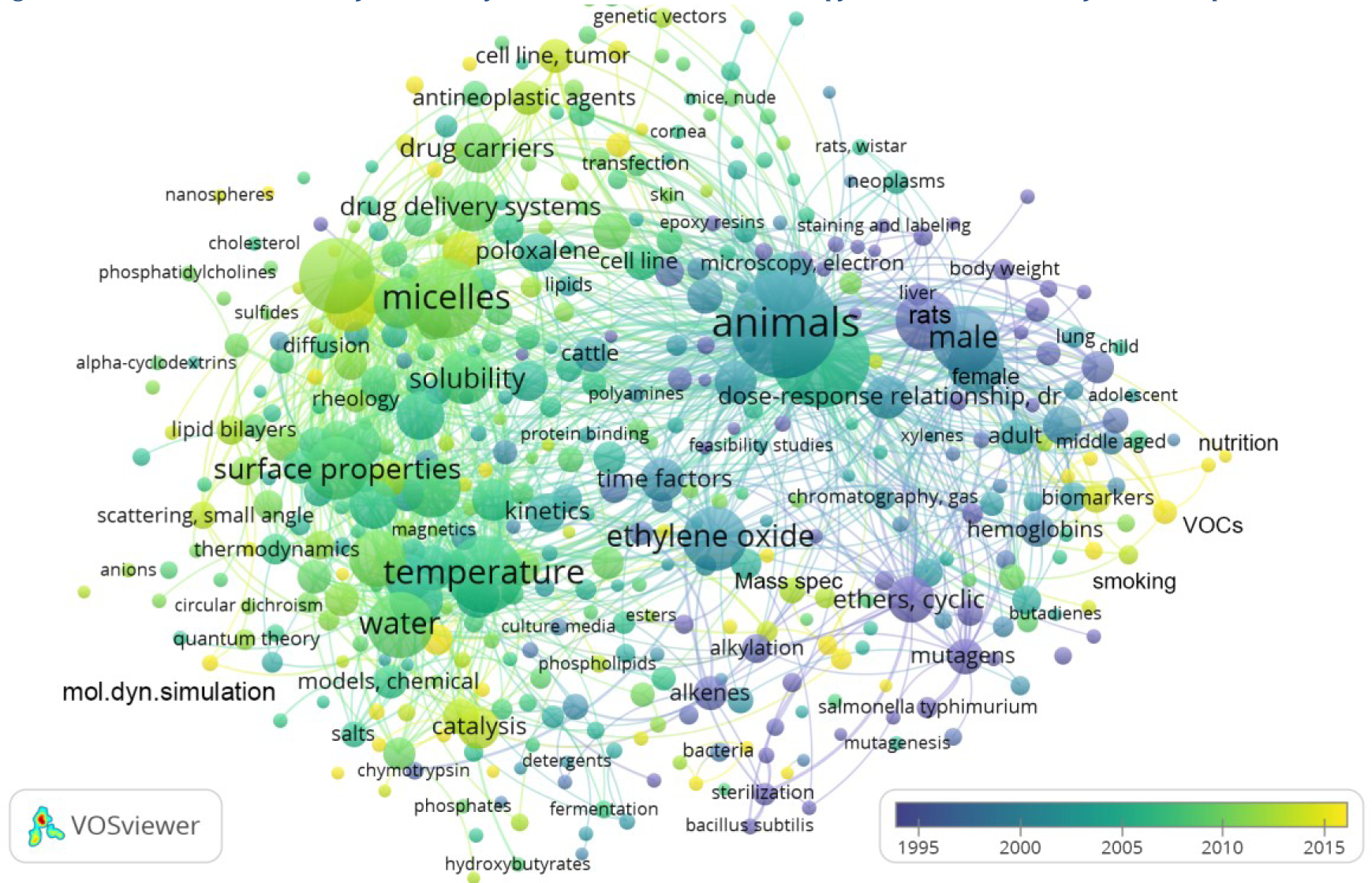
VOSviewer timeline of MESH Keywords Associated with Propylene Oxide Articles from EuropePMC

Even while studies connecting tobacco use and propylene oxide have decreased, the surge in e-cigarettes prompted investigations into toxic emissions from vaping products. The most cited article in our collection (Sleiman et al., 2016) highlighted that propylene oxide remained a key component in e-liquid refills. An earlier study observed the formation of thermal degradation byproducts during vapor production, noting that when heated and vaporized, propylene glycol can form propylene oxide (Grana et al., 2014).

As of 2014, propylene oxide has been reevaluated as a potential biomarker for breast cancer (Turner et al. 2023, Garcia et al. 2015). Recent research, exemplified by papers such as (Pal & Kannan, 2023, Li et al., 2023, Mendy et al. 2022, and Liu et al., 2022), has consistently illuminated the implications of propylene oxide exposures in both humans and animals. These studies indicate associations with various health aspects, including pregnancy and IVF treatment, reduced lung function, oxidative stress, cancer, diabetes, and neurological conditions like dyslexia, highlighting that the broader population’s exposure to propylene oxide is indeed a public health concern.

## Summary of Included Studies

The systematic review’s literature search, outlined in the methodology and illustrated in the PRISMA diagram (Figure 2), identified over 10,000 records from diverse sources.

**Figure 2.**
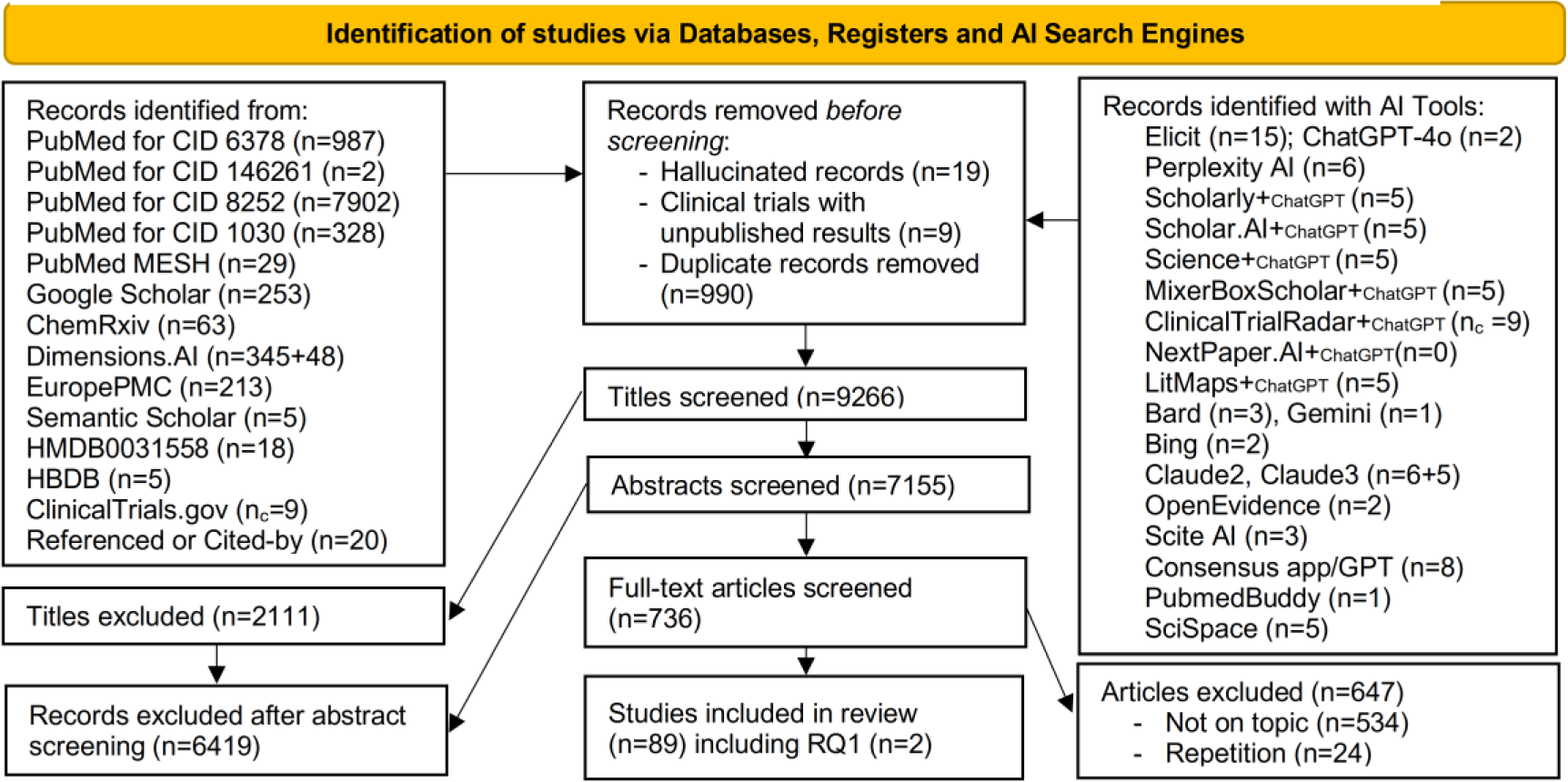
PRISMA flow diagram

The search strategy encompassed databases such as PubMed, Google Scholar, ChemRxiv, Dimensions.AI, Semantic Scholar, and the Human Metabolome Database (HMDB), among others, with PubMed yielding the majority of records, particularly for PubChem CID 8252 (Propylene), even though indexing has not extended beyond 2012. Additionally, our review encompassed records uncovered through AI tools, including various ChatGPT integrations and standalone platforms, which contributed additional unique records. Moreover, several specialized databases were queried. Notably, the Human Metabolome Database (HMDB) features a comprehensive entry for Propylene Oxide (Methyloxirane, entry HMDB0031558). While PO’s presence is “Expected but not Quantified” in biological specimens, the database includes 18 references that were integrated into the search. Further exploration via the Human Breathomics Database, while not directly providing information on PO levels, offered insights into compounds structurally or functionally akin to Propylene, underscoring the nuanced approach necessary to compile scattered yet pertinent data pieces.

Searches were conducted multiple times between April, July, and November 2023, with final searches in January and February 2024, and an additional search for this revision in May 2024. Results from PubChem, PubMed, HMDB, and HBDB remained consistent throughout this period. The HAWC resource was last updated in October 2023. The most notable changes were observed in Dimensions.AI, which provided the most up-to-date sources, and Google Scholar. While ChemRxiv publications showed constant growth, they were less relevant to the research questions of this review.

The extensive screening process began with the elimination of irrelevant or redundant records, including hallucinated entries, unpublished clinical trials, and duplicates, culminating in a focused review of 9,266 titles. Title screening further narrowed down to 7,155 abstracts, and eventually, 736 full-text articles were scrutinized for relevance and quality. 24 articles had relevant information but were excluded as providing repeated or non-original information or interpretation available in newer sources. Ultimately, 89 articles were deemed pertinent to our study’s objectives, with a single article directly addressing our primary research question (RQ1), while the remainder contributed insights relevant to secondary questions (RQ2-RQ6).

**Table 1.** summarizes compounds included in literature search for this review.

| Compound | Relation to Propylene Oxide |
| --- | --- |
| Propylene also known as Propene (PE) | Upstream (Precursor) |
| 1,3-propanediol (PDO) | Upstream (Precursor) |
| Propylene Glycol (PG) | Downstream (Metabolite), but could be also Upstream (Precursor) |
| 1-Methoxypropan-2-yl Acetate | Downstream (Derivative) |
| 1-(1-Methoxypropan-2-yloxy)propan-2-ol | Downstream (Derivative) |
| 1,2-propanediol | Downstream (Derivative) |
| N-Acetyl-S-(2-hydroxypropyl)-L-cystein (2HPMA) | Downstream (Metabolite, Urine Biomarker) |
| 3-HP (3-hydroxypropionaldehyde) | Downstream (Metabolite, Blood Biomarker) |
| 7-Hydroxypropylguanine (7-HP-guanine), 3-HP-adenine, 1-HP-adenine, 3-HP-cytosine | Downstream (DNA adducts, Blood Biomarkers) |
| N-(2-hydroxypropyl)valine (HOPrVal) | Downstream (Hemoglobin Adduct, Blood) |
| N-(2-hydroxypropyl) histidine (N-(2-HP) histidine) | Downstream (Metabolite, Blood Biomarker) |

Information related to these compounds was considered to gain potential insights into the metabolic and environmental pathways that could influence the presence and metabolism of Propylene Oxide in the human body.

In our research on propylene oxide, we utilized the [Public Assessment of Propylene Oxide](https://hawcproject.iarc.who.int/lit/assessment/705/) hosted by Health Assessment Workspace Collaborative (HAWC) platform, following its mention in the latest IARC review on Propylene (Turner et al. 2023) – one of papers in our final list of included studies.

HAWC serves as a comprehensive content management system aimed at storing, displaying, and synthesizing environmental and human health assessment data from diverse sources. It supports the systematic review process, a cornerstone of organizations such as the International Agency for Research on Cancer (IARC) and the United States National Toxicology Program (NTP), by providing a platform for detailed literature search, data extraction, and analysis. HAWC organizes data into assessments, collaborative projects that facilitate the extraction and visualization of data from peer-reviewed publications. This process is streamlined through specialized data extraction forms and workflows.

Systematic review is a historical strength of the International Agency for Research on Cancer (IARC) Monographs Program and the United States National Toxicology Program (NTP) Office of the Report on Carcinogens (RoC). Both organizations are tasked with evaluating peer-reviewed, published evidence to determine whether specific substances, exposure scenarios, or mixtures pose a cancer hazard to humans. This evidence synthesis is based on objective, transparent, published methods that call for extracting and interpreting data in a systematic manner from multiple domains, including a) human exposure, b) epidemiological evidence, c) evidence from experimental animals, and d) mechanistic evidence. The process involves multiple collaborators and requires an extensive literature search, review, and synthesis of the evidence.

The [Public Assessment of Propylene Oxide](https://hawcproject.iarc.who.int/lit/assessment/705/) on HAWC, initiated by IARC in 2022 and updated in 2023, focused on evaluating the carcinogenicity and genotoxicity of PO. This assessment incorporated findings from a systematic literature review, covering various aspects from identification and exposure to carcinogenicity and genotoxic effects of PO. Notably, it included sections on oxidative stress, genotoxicity, and epigenetics, among others, contributing comprehensive insights into PO’s impact. Propylene Oxide was considered by previous Working Groups, in February 1976, June 1984 and March 1987 (IARC, 1987). Newer data was incorporated into the 1994 monograph and the latest 2023 monograph (Turner et al. 2023) reported that since the last IARC review in 1994, only one epidemiologic study of U.S. propylene oxide manufacturing workers has been published; the authors did not find increased mortality due to cancer by duration of exposure with or without latency, nor did they find increased cancer risk by process. Recent exposure and biomarker studies have shown that PO forms chemically stable hemoglobin and DNA adducts and that concentrations of these adducts are related linearly to air concentrations of PO. In addition, hemoglobin and DNA adducts and sister chromatic exchanges were reported

The [Public Assessment of Propylene Oxide](https://hawcproject.iarc.who.int/lit/assessment/705/) includes 675 references related to the compound’s environmental and health impacts. Out of our final selection of 89 articles, 43 were categorized within HAWC, with the remaining 46 found outside the platform.

After review of included studies, we changed RQ2 from “How do health conditions affect the levels of propylene oxide detected in exhaled breath? to RQ2* “How different concentrations of PO, its precursors and derivatives are associated with various health conditions”, since there was no research addressing the original question. The table below (Table 2) summarizes how each RQ of this review aligns with the HAWC Public Assessment sections.

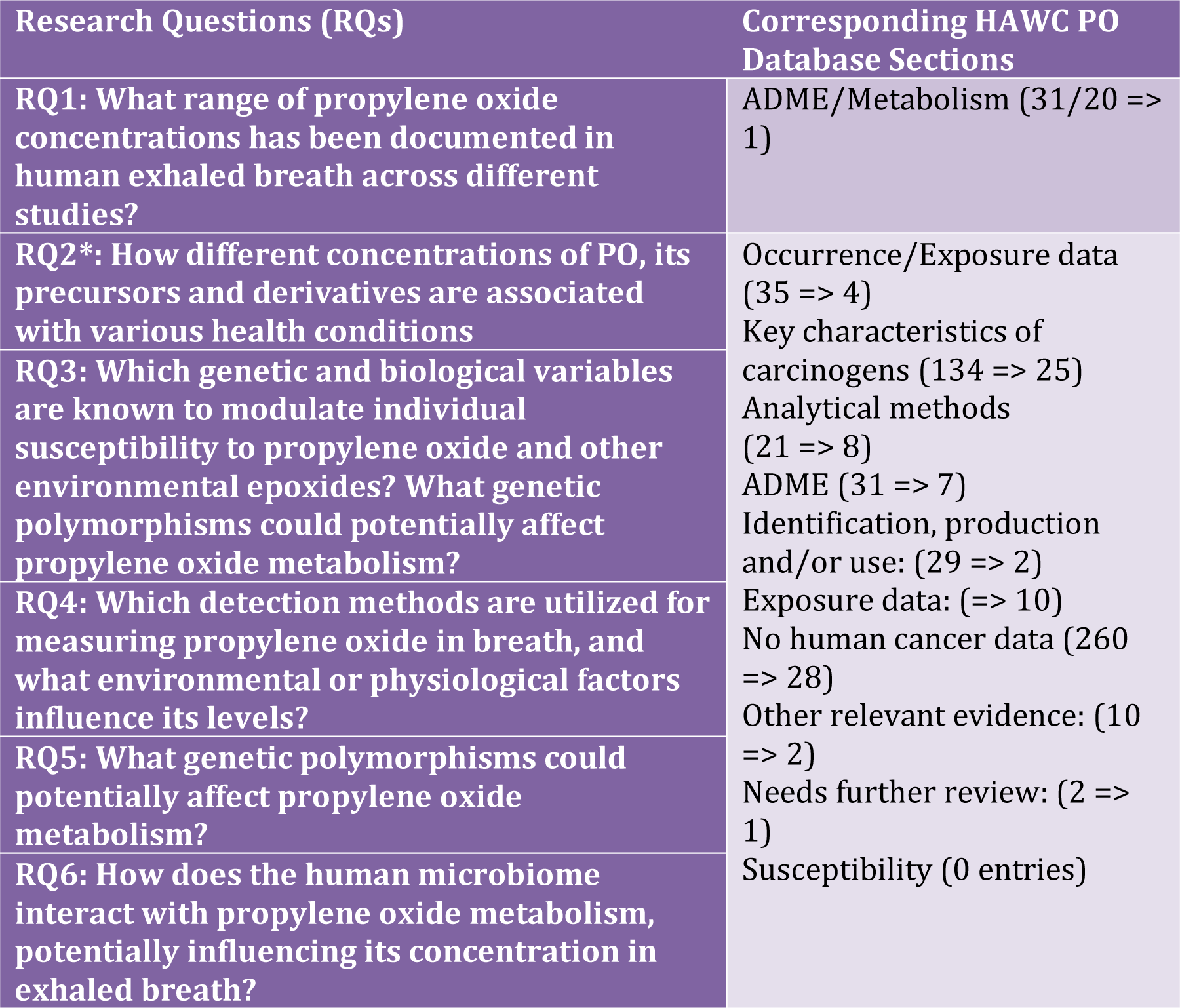

We note that “susceptibility” is included among the categories in the “other adverse effects” section, yet it notably lacks citations. This gap underscores the need for studies focused on understanding the variables that affect PO metabolism and their consequent implications for human health.

### Propylene Oxide in Exhaled Breath

Despite conducting an extensive literature search, we found that studies directly addressing propylene oxide concentrations in exhaled breath are remarkably scarce. The Human Breathomics Database (HBDB), which includes compounds with a propylene (propan-2-yl) group, lacks specific information on propylene oxide. Similarly, the Volatilome List of the [Comptox database](https://comptox.epa.gov/dashboard/chemical-lists/VOLATILOME), which compiles compounds detected in human breath and reported in peer-reviewed literature and experimental work at the US-EPA, does not include any data on propylene oxide, featuring only 1-propene and [Propylene Glycol](https://comptox.epa.gov/dashboard/chemical/details/DTXSID0021206). The Human Metabolome Database (HMDB) indicates that propylene oxide concentrations in biological specimens are not properly quantified. This gap underscores the challenge in establishing standard concentration ranges and dependencies for propylene oxide in exhaled breath.

Despite conducting an extensive literature search, we found that studies directly addressing propylene oxide concentrations in exhaled breath are remarkably scarce. The Human Breathomics Database (HBDB), despite including compounds with a propylene (propan-2-yl) group, lacks information on propylene oxide. The Volatilome List of [Comptox database](https://comptox.epa.gov/dashboard/chemical-lists/VOLATILOME) - a subset of compounds detected in human breath and reported in the peer-reviewed literature and identified in experimental work at US-EPA - does not have any information on Propylene Oxide and features only [Propylene](https://comptox.epa.gov/dashboard/chemical/details/DTXSID5021205?list=VOLATILOME) and [Propylene Glycol](https://comptox.epa.gov/dashboard/chemical/details/DTXSID0021206). HMDB reports that Propylene Oxide concentrations in biological specimens are not properly quantified. This gap underscores the challenge in establishing standard concentration ranges and dependencies for PO in exhaled breath.

A singular reference emerged as the only one directly answering this main question of this systematic review (RQ1) – a team of toxicologists from Germany (Filser et al. 2008) that first reported the PO was detected in human breath after exposure to Propylene in an earlier 1997 study (not retrievable from online literature databases) published in German in Naunyn–Schmiedeberg’s Archives of Pharmacology, the official journal of the German Society of Experimental and Clinical Pharmacology and Toxicology (DGPT), founded in 1873. It is the oldest journal in this field of science. The 2008 study further expanded this research to careful examination of exposures in 4 volunteers and laboratory animals.

This investigation by Filser et al. (2008) stands as the only study identified through a PubMed search combining “propylene oxide” and “exhaled breath”. Interestingly, this reference was also frequently identified by AI search tools such as Elicit and Perplexity in response to the main query of this paper, though not universally across all platforms (notably absent in MixerBoxScholar). It ranked highly on Semantic Scholar. These observations suggest that while AI tools can pinpoint precise answers to specific research questions, their efficacy diminishes when seeking nuanced, indirectly relevant information in the absence of direct answers, a topic we explore further in the subsequent sections.

The Filser study documented PO concentrations in human exhaled air ranging from 0.083 ppb to 0.3 ppb, following exposures to 10 ppm and 25 ppm of propylene for 180 minutes. The study provided graphical representations of PO concentration-time courses in exhaled air and discussed blood concentrations predominantly, with brief mentions of exhaled air concentrations. It estimated that 6.6% to 11.9% of inhaled propylene was metabolized to PO, indicating dose-dependent metabolic conversion with metabolic rates between 8.57 µmol/h and 38.1 µmol/h. Obviously, there was a considerable difference between individuals and it did not correlate with age or body mass. However, the discussion primarily focused on the liner relationship between blood PO and air PE concentrations.

A significant finding was the pronounced interspecies differences in PO metabolism, highlighting substantial variances in the activities of PO-metabolizing enzymes between humans and rats. This suggests a species-specific first-pass metabolism of epoxides, metabolically derived from olefins, within the liver.

The study concluded that the low PO concentrations observed in humans exposed to PE necessitate further research to elucidate the mechanisms or toxicokinetics involved. The inferred health risk from PE exposure, via metabolically produced PO, appeared to be exceedingly low, supported by the significantly lower blood concentrations of metabolically derived PO compared to those from direct inhalation.

The detection of epoxides like PO in exhaled breath, traditionally considered rare, may reflect environmental exposures rather than direct disease associations. However, the findings also suggest individual metabolic variations. The Filser et al. (2008) study, as the only one reporting PO concentrations in exhaled breath, highlighted considerable individual variability in these concentrations, independent of body mass or respiration rates, underscoring the need for further studies to elucidate PO’s biotransformation within the human body.

Our newest study [5] now adds a second reference directly answering RQ1. Correctly identified by the newest version of Gemini and GPT4-o, it is also highly ranked by SemanticScholar, Dimensions.AI, EuropePMC, and GoogleScholar. Our study shows high levels of PO in exhaled air concentrations in a subset of environmentally sensitive individuals likely having less efficient enzymes unable to detoxify the compound or altered mechanisms of absorption, delaying release of this pollutant or byproduct of other pollutants such as Propene. Our results are in line with other studies that identified byproducts of PO in blood and urine [25, 26], showing that exposure to this pollutant is higher than previously thought. Our study aligns with Filser’s work also showing that individual metabolism makes a big difference.

### Propylene Oxide Exposures

Although Propylene Oxide is discernible by odor, its scent can vary considerably among individuals, ranging from fruity and floral to pungent, medicinal, or solvent-like. Additionally, the threshold at which its odor becomes detectable varies significantly, spanning from 10 to 200 ppm depending on individual sensitivity. Level of Distinct Odor Awareness (LOA) to PO is set to 21 ppm by US EPA.

Exposure to **Propylene Oxide** in occupational settings varies, with manufacturing sites often maintaining levels below 0.1 to 5 parts per million (ppm) due to effective control measures (Czene et al., 2002, Ogawa et al. 2006). However, certain environments report concentrations ranging from 1 to 7 ppm, correlating with elevated biomarkers of exposure, such as DNA and hemoglobin adducts, indicating its genotoxic potential. It is important to note that the concentrations of propylene oxide found in breath may not be directly proportional to the concentration of propylene oxide in the air, as other factors such as ventilation and individual differences in metabolism can affect the amount of propylene oxide that is absorbed and exhaled.

PO is mass-produced, contributing to its ubiquity. The routes of exposure to propylene oxide are inhalation, ingestion, and incidental dermal exposure. Consumers may be exposed through ingestion of propylene oxide residues in foods resulting from its use as an indirect food additive or by contact with consumer products containing propylene oxide. The U.S. Environmental Protection Agency (EPA) has established tolerance limits for propylene oxide residues from fumigation of cocoa beans, nutmeats, herbs, spices, and some fruits (e.g., figs, prunes, and raisins). Consumer products with the highest concentrations of propylene oxide include automotive and paint products, with some automotive products listing concentrations of 0.1% to 0.5% [35]. Consumer products with the highest concentrations of propylene oxide include automotive and paint products, with some automotive products listing concentrations of 0.1% to 0.5%.

Exposure to PO can also occur through items such as cellulose acetate film, wood shavings, paper cups, rubber tubing, nylon film, polycarbonate film, and polyvinylidene chloride film may contain residual PO from sterilization processes, use of sterilized products like wrappings, tubings, medical equipment, and food items may contain residual PO or ethylene oxide, leading to exposure when used. Despite efforts to eliminate gas residues through aeration, accidental exposure to residual gas in sterilized items still occurs. Workers involved in sterilizing items or handling sterilized products may be exposed to PO. This exposure can extend to family members through contact with clothing emitting the chemical. Additionally, the formation of PO in alkali metal batteries [44] poses another potential route of exposure, especially for individuals working in battery manufacturing, handling, or recycling.

Foods treated with PO as a fumigant, such as cocoa, gums, processed spices, starch, and processed nut meats, can contain residues of the chemical. PO is utilized by the pistachio and almond industry to achieve a 5-log reduction of Salmonella Enteritidis, serving as a fumigant to reduce bacterial, mold, and yeast populations [40]. For almond and walnut fumigation, the daily time-weighted-average exposure concentration was 0.71 ppm (geometric mean) for combined non-specific exposure and exposure during chamber unloading (adjusted for exposure duration) [35]. It is often assumed that consumers are not exposed, as PO almost immediately dissipates and breaks down after application, but this has not been sufficiently studied. Cao and Corriveau [3] examined 36 selected food composite samples from the 2007 Canadian total diet study. Propylene oxide was not detected in any samples analyzed with an average method detection limit of 0.5 ppb/g. This finding suggested that propylene oxide is very unlikely to be found in foods as consumed due to its volatility and reactivity with water.

Propylene oxide is a volatile organic compound (VOC) that breaks down relatively quickly in the environment. It has an atmospheric half-life of 3 to 20 days and is also water-soluble, which means that it can be washed away by precipitation.

Interest in the health effects of PO exposure in the general population was limited until recently. The US Environmental Protection Agency and the European Union have focused mainly on acute and chronic occupational exposure. The European Union’s Risk Assessment Report concluded that exposures to PO in local and regional scenarios are extremely low, hence the degree of risk is anticipated to be negligible. However, since environmental, air, food, and water contamination all contribute to overall PO exposure (Dauchet, 2023), it might not fully account for the observed associations in studies. Other environmental exposure sources include tobacco smoke, with University of Kentucky Reference Cigarettes 1R4F and 2R4F containing 0.93 and 0.65 microg/cigarette of PO, respectively (Diekmann et al. 2006). Additionally, functional fluids (such as hydraulic fluids, heat transfer fluids, and lubricants), propylene oxide–based surfactants, soil sterilizer, fumigants in agricultural products and contaminants in air, food, and water are identified as significant contributors to overall PO exposure.

**Propylene Glycol** (PG) in cigarettes and e-liquids can also be converted to PO, contributing to exposure (Grana et al., 2014). PG is used in various products, including personal care (hair products, toothpaste and cosmetics), cleaning, and water-based paints (Wieslander and Norbäck, 2010) as well as a food additive (as an antifreeze in ice cream), in pharmaceutical preparations, in cosmetics, and in workplace environments like smoke generators in discotheques, theatres, and aviation emergency training (Wieslander et al. 2001).

**Propylene**, a precursor to Propylene Oxide, is detectable even in remote locations, sourced from diverse emissions such as biomass burning and fossil fuel combustion (Morgott, 2018). It is also detected in interstellar space and celestial atmospheres (Singh et al., 2022), indicating its widespread presence independent of human activities. In central Taiwan, daily average values range from 0.3 to 6.7 ppb, likely due to traffic-related sources (Tsai et al., 2010). Texas records propylene levels from various sources, including industrial emissions and vehicle exhaust, with the highest annual average air concentration being 7.9 ppbv in Houston (Myers et al., 2015).

Indoor propylene exposure is typically 2-3 times higher than outdoors due to residential activities like cooking and heating (Morgott, 2018). Exposure is even higher in smoking households. Urban areas generally have more variable and higher concentrations (up to 10 ppbv) than rural regions (< 2ppbv). Elevated PE exposures may occur in specific workplaces. Firefighters or refinery plant operators may encounter levels up to about 10 ppmv.

**1,3-Propanediol** (PDO), used in industrial polyesters and found in municipal refuse incineration stack effluents, is naturally generated through microbial fermentation in foods and beverages like sauerkraut and wine. PDO is frequently employed in cosmetics and can be oxidized to propylene oxide. While theoretical pathways suggest PDO conversion in the human body, evidence is less clear, but we included two records on PDO in our final list of selected publications.

## Methods for Quantification

To directly measure levels of PO in exhaled breath, techniques such as Gas Chromatography-Mass Spectrometry (GC-MS), Selected Ion Flow Tube Mass Spectrometry (SIFT-MS), Proton Transfer Reaction Mass Spectrometry (PTR-MS) and Ion Mobility Spectrometry (IMS) can be employed. Currently, there are no commercially available e-noses or breathalyzers specifically designed to measure PO in air or breath due to challenges in low-concentration detection, specificity and sensitivity, as well as high R&D costs required.

In the top study selected for this review (Filser et al. 2008), PO was measured using gas chromatography with flame ionization detection (GC/FID) for animal experiments and gas chromatography with mass-selective detection (GC/MSD). A method based on isotope dilution headspace and GS-MS was developed for the determination of propylene oxide in foods [3]. GS-MS was also used by Ball et al. 2005 and Diekmann et al [9] while Schettgen et al [42] utilized liquid chromatography/tandem mass spectrometry for simultaneous quantification of five mercapturic acids derived from several industrial chemicals including PO. The solvation of propylene oxide in water has been studied using vibrational circular dichroism (VCD) spectroscopy, optical rotation dispersion (ORD) spectroscopy, molecular dynamics simulations, and ab initio calculations. Immunoassays (such as enzyme-linked immunosorbent assays, ELISA, and radio-immunoassays), Edman degradation and enzymatic assays (eg, Epoxide Hydrolase and Glutathione S-Transferase Assays) were also among the methods used.

In addition to direct measurements of propylene oxide and biological markers of exposure, computational modeling has been employed to quantify PO exposure. For instance, Csanády and Filser (2007) developed physiological toxicokinetic models for PO in rats and humans, aiding in risk estimation. Furthermore, Oldham et al. (2021) utilized computational fluid dynamics to forecast passive exposure across diverse environments, enriching our understanding of exposure dynamics.

Computational statistical models have also been utilized to assess the risk of health effects, as demonstrated in studies such as the California Teachers Study, which involved women diagnosed with invasive breast cancer (Garcia et al. 2015). Here, estimated ambient concentrations of hazardous air pollutants (HAPs), including PO, were computed using EPA assessments. Annual average ambient air concentrations of 24 mammary gland carcinogens (MGCs), including PO, were linked to participants’ addresses. Cox proportional hazards models were then employed to estimate hazard rate ratios and associated confidence intervals for residential MGC levels. MGCs were analyzed both individually and as a combined summary variable for all participants, in selected subsets, and by tumor hormone responsiveness.

### Health Effects

The EPA produces the National-Scale Air Toxics Assessment (NATA) to identify and prioritize air toxics with respect to their potential population health risk. The results from the [AEGL Program of the US EPA for PO](https://www.epa.gov/aegl/propylene-oxide-results-aegl-program) indicate the following thresholds of exposure:

AEGL-1: At 73 ppm, individuals in the general population, including those who are susceptible, may experience notable discomfort, irritation, or certain asymptomatic, non-sensory effects. These effects are not disabling and are transient, reversing upon cessation of exposure.

AEGL-2: Exposure to propylene oxide at concentrations ranging from 440 ppm for 10 minutes to 86 ppm for 8 hours may lead to irreversible or other serious, long-lasting adverse health effects, or impair an individual’s ability to escape.

AEGL-3: Exposure to 1300 ppm of propylene oxide for 10 minutes could result in life-threatening health effects or death.

The table provides a summary of the currently accepted values in ppm [32].

| Propylene oxide |  | CAS #:75-56-9 |  |  |  |
| --- | --- | --- | --- | --- | --- |
|  |  | ppm |  |  |  |
|  | 10 min | 30 min | 60 min | 4 hr | 8 hr |
| AEGL 1 | 73 | 73 | 73 | 73 | 73 |
| AEGL 2 | 440 | 440 | 290 | 130 | 86 |
| AEGL 3 | 1300 | 1300 | 870 | 390 | 260 |

The health effects of PO exposure have been extensively studied across various models, including in vitro systems, animal studies, and observational studies in human populations. These studies collectively indicate that PO has the potential for causing a range of acute and chronic conditions, from cellular and genetic damage to carcinogenic outcomes, depending on the concentration of exposure and the duration. Studies also indicate associations with reduced lung function, oxidative stress, hyperglycemia, diabetes, nervous system effects, peripheral nerve damage, potential neurotoxicity and neurological conditions like dyslexia in children [8].

At the molecular level, PO forms adducts with DNA and hemoglobin, serving as biomarkers of exposure. The presence of N-(3-hydroxypropyl) valine (HOPrVal) in workers’ blood samples correlates with workplace exposure levels. Epidemiological studies have also linked PO exposure to cytogenetic effects, including increased levels of DNA adducts and sister chromatid exchanges in exposed workers. Smokers, in particular, exhibit higher levels of urinary 2-HPMA, indicative of PO’s contribution to toxicant exposure.

The toxicity of propylene oxide has been studied in several animal species. Acute oral median lethal dose (LD50) values of 380 mg kg−1 in the rat and 660 mg kg−1 in the guinea pig have been reported. The 4 h median lethal concentration (LC50) value for inhalation is 4000 ppm for the rat and 1740 ppm for the mouse. Propylene oxide neuropathy has been demonstrated in rats. In a 7-week study, animals exposed to propylene oxide at a concentration of 1500 ppm in air, developed ataxia in the hind legs.

PO has exhibited mutagenic properties in bacterial and mammalian systems in vitro. However, there is limited evidence of mutagenicity in vivo, except for specific instances such as an increase in micronucleated erythrocytes in mice subjected to intraperitoneal exposure to a high dose (300 mg/kg body weight) of the compound, and an increase in sister chromatid exchanges observed in mouse bone marrow cells at intraperitoneal doses of 300 and 450 mg/kg body weight.

Inhalation studies on monkeys exposed to 100 or 300 ppm of PO (6 hours/day, 5 days/week for 104 weeks) did not show a significant increase in sister chromatid exchanges or chromatid-type aberrations in peripheral lymphocytes when compared to control groups (as reviewed in Rias Blanco et al., 2002, and references therein). Additionally, exposure-dependent accumulation of N-(2-hydroxypropyl)valine has been observed in the hemoglobin of F344 rats exposed to propylene oxide via inhalation.

In F344 rats exposed to 400 ppm propylene oxide for 6 hours per day, 5 days per week for up to 103 weeks, a low incidence of nasal papillary adenomas (2 out of 50 males, 3 out of 50 females) was observed. Similarly, in mice subjected to chronic exposure to 400 ppm propylene oxide, there was a low occurrence of nasal hemangiomas and hemangiosarcomas (Renne et al., 1986). No increases in tumor incidence were noted for the groups exposed to 200 ppm. These findings align with those reported by Lynch et al. (1984), where two adenomas were observed in the nasal passages of 80 male F344 rats exposed to 300 ppm propylene oxide for 7 hours per day, 5 days per week for 104 weeks; however, no tumors were observed in the group exposed to 100 ppm (80 male F344 rats). Wistar rats exposed to 0, 30, 100, or 300 ppm propylene oxide for 6 hours per day, 5 days per week for 123–124 weeks did not show a significant increase in nasal tumors [10]. However, continuous, severe perturbation of GSH in RNM following repeated high PO exposures led to inflammatory lesions and cell proliferation, which could in their turn lead to tumorigenicity (Lee et al. 2004).

Non-neoplastic nasal lesions following chronic propylene oxide exposure include respiratory epithelial cell hyperplasia, squamous metaplasia, and olfactory epithelial degeneration. Notably, some non-neoplastic histopathological effects were observed at exposure concentrations lower than those associated with tumor formation [10].

Urinary metabolites of propylene oxide were associated with lower lung function independently of smoking (Liu et al. 2022).

Lynch et al. (1984) exposed groups of 12 cynomologus monkeys to 0 (filtered air), 100, of 300 ppm of propylene oxide for 7 hr/day, 5 days/week over a 2-year period. Blood was collected during the final month of exposure and used to culture lymphocytes to assay sister chromatid exchanges (SCEs) and chromosomal aberrations. The incidence of SCEs; or chromosomal aberrations was not significantly altered compared with the control group.

The table below presents a sample of heterogeneous experiments, observations, and conditions, demonstrating the complexity in the integration and synthesis of the data.

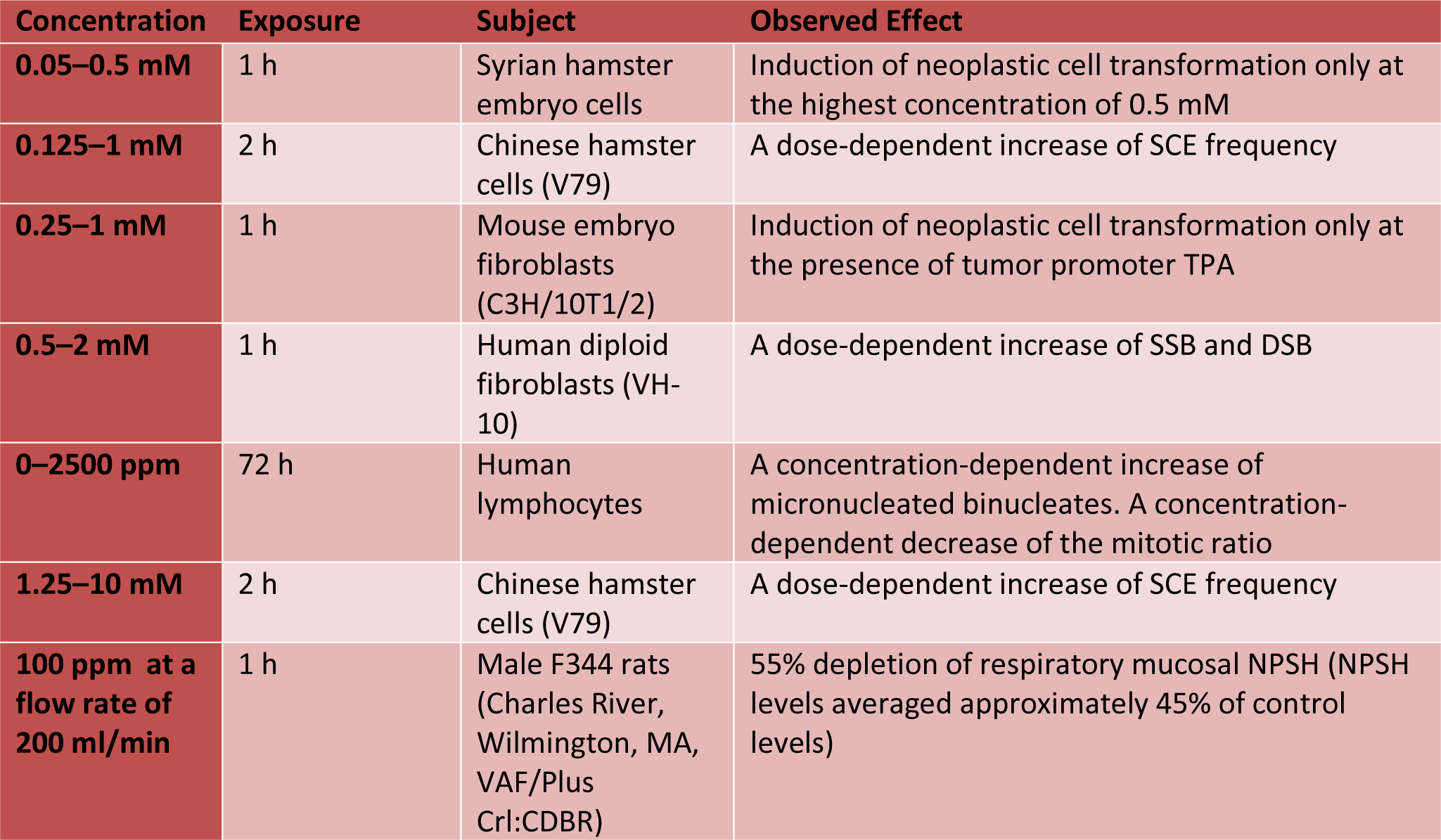
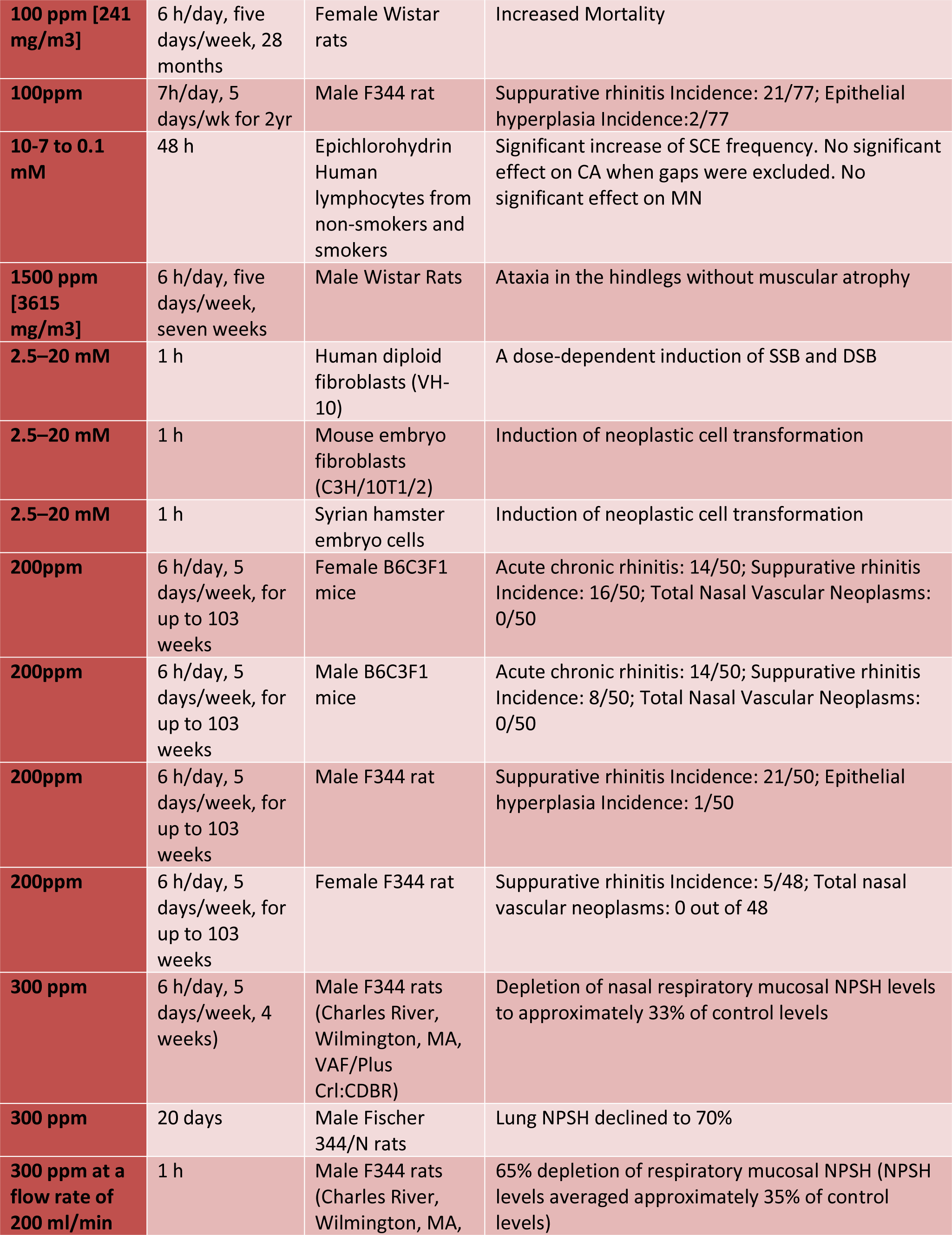
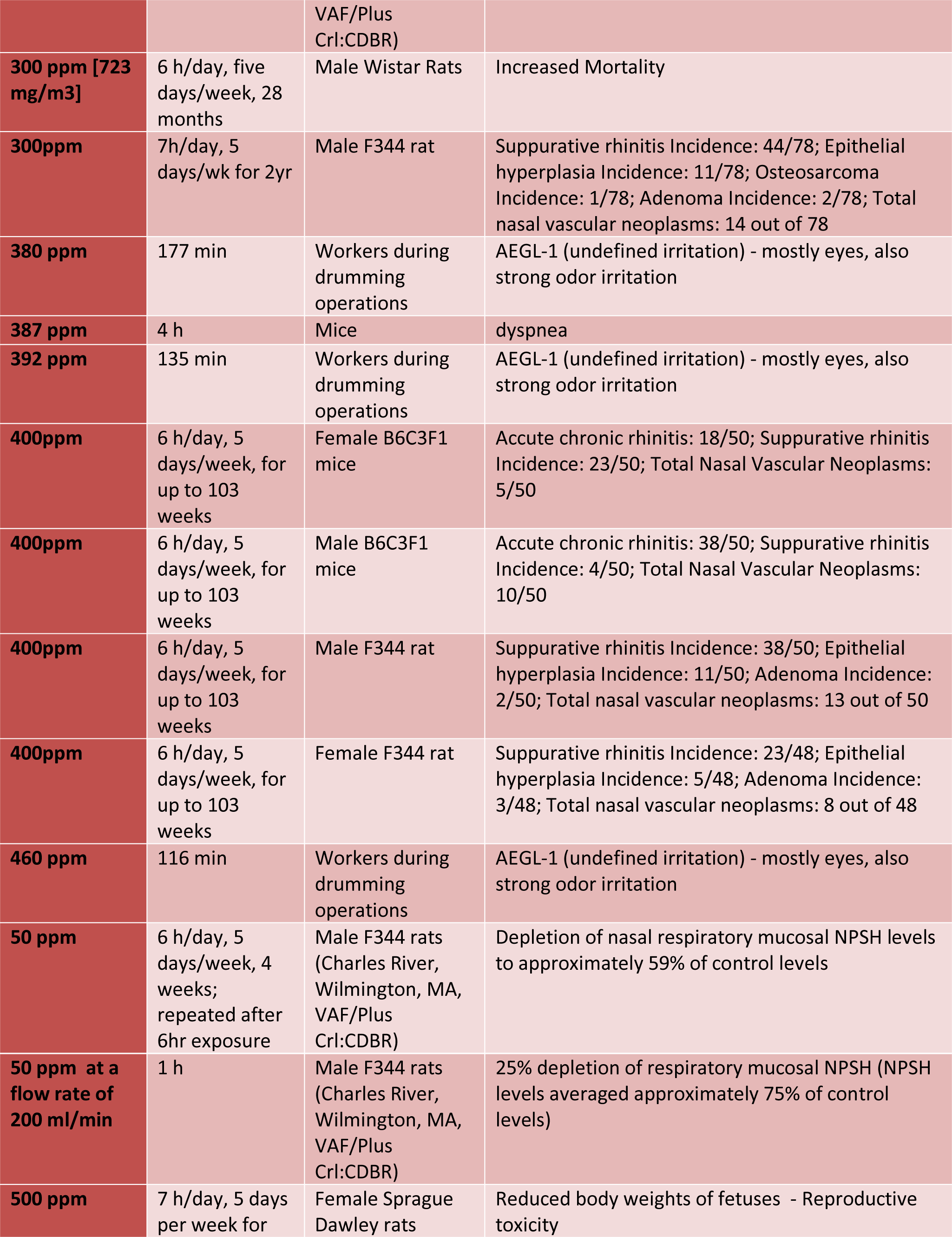
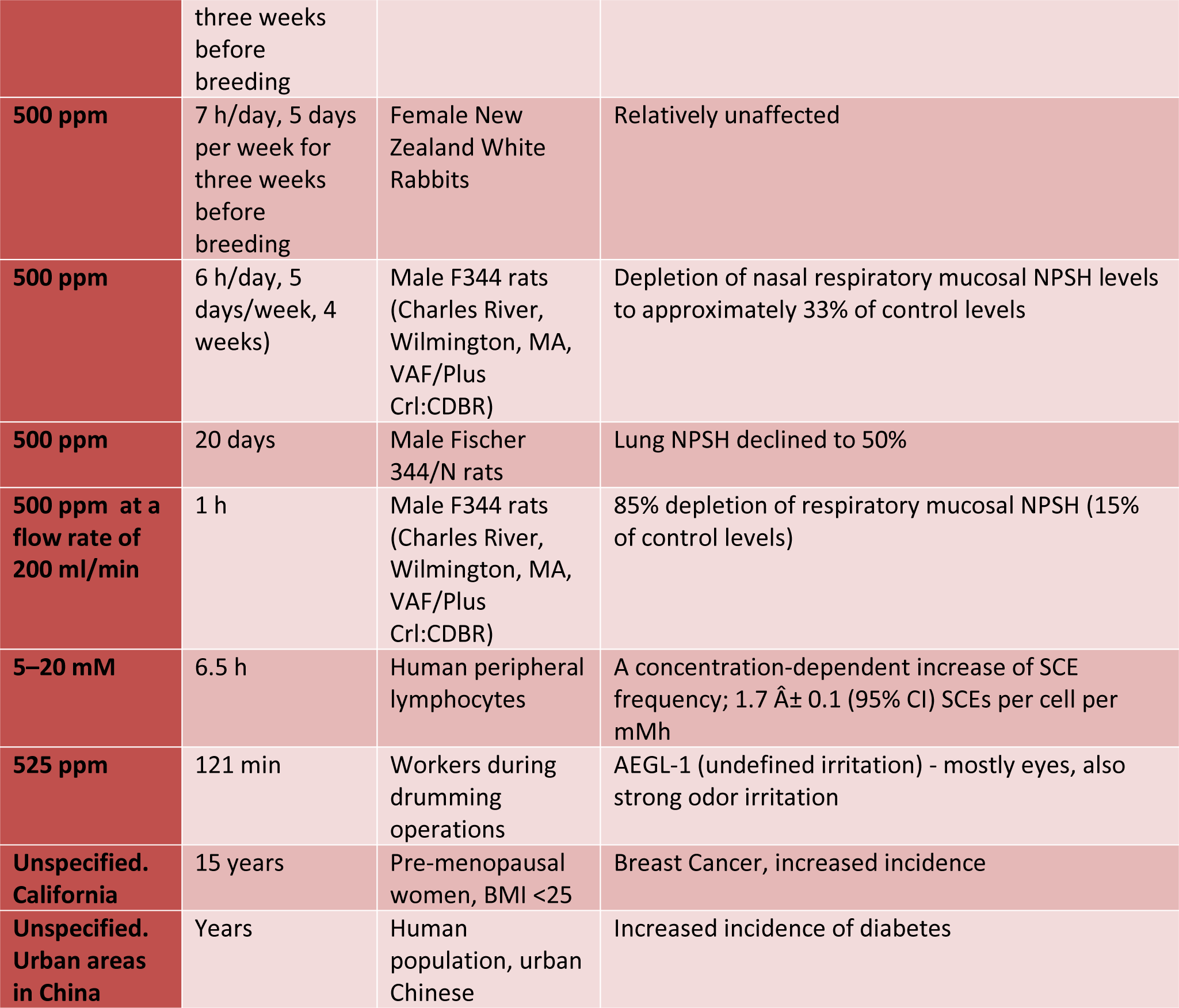

Studies on toxicity of possible PO precursor PDO yield contradictory results, with severe health effects reported in individuals at high concentrations, yet inhalation studies in rats show no adverse effects up to 1800 mg/m³ (Garg et al., 2008; Scott et al., 2005).

Propylene Glycol, a metabolite and possible precursor of PO, is tolerated well in pharmaceutical preparations [Krakoff et al. 2001], but there are multiple Case Reports and Clinical Observations of negative effects.

PG from smoke can lead to acute ocular and respiratory effects [Wieslander et al. 2001). Short exposure to PG mist from artificial smoke generators may cause acute ocular and upper airway irritation in non-asthmatic subjects. A few may also react with cough and slight airway obstruction.

Propylene glycol (PG) exposure, particularly in high doses or prolonged administration, can lead to metabolic acidosis, hyperosmolarity, and hyperlactatemia. This can occur due to the accumulation of PG in the body, especially in individuals with compromised renal function induced by conditions like cocaine use. Metabolic acidosis and hyperlactatemia were observed in a patient who received a high dose of lorazepam, which contains propylene glycol, over 72 hours.

PG has been implicated in neurotoxicity, particularly in children. Studies have shown that PG exposure can trigger widespread apoptotic neurodegeneration in the developing central nervous system, especially at certain developmental stages. Additionally, when combined with certain medications like phenobarbital, which also induce apoptosis in the immature central nervous system, PG can exacerbate neurotoxicity.

Skin Irritation and Allergic Reactions: PG may cause allergic or irritant contact dermatitis in some individuals. Patch-test studies have demonstrated that a small percentage of patients exhibit allergic reactions to PG, particularly when it is present in personal care products, topical medications, or occupational settings.

Studies in neonates have highlighted potential renal and hepatic toxicity associated with prolonged exposure to PG, particularly in high concentrations. However, short-term exposure to regulated concentrations of PG in neonates did not result in adverse effects on renal, metabolic, or hepatic function.

Preclinical studies in rats and dogs demonstrated that aerosolized cyclosporine formulated in propylene glycol did not result in respiratory or systemic toxicity at exposures up to 2.7 times the maximum human exposure. However, the effects of aerosolized PG on respiratory and systemic health may vary depending on factors such as exposure concentration and duration. Negative health effects could be exacerbated in children and those with compromised renal function.

Several case studies highlight acute toxicities associated with propylene glycol (PG), a compound related to PO. For instance, acute PG intoxication in children has been observed, resulting in metabolic acidosis and systemic toxicity due to high exposure levels, as seen in a two-year-old experiencing lethargy and polypnea after ingesting disposable cleansing towels containing PG. Similarly, neurotoxic effects of PG in pediatric medicine have been reported, emphasizing its potential apoptotic impact on the developing central nervous system.

In veterinary medicine, a case of propylene glycol intoxication in a dog (Claus et al. 2011) demonstrates the compound’s systemic effects and the successful use of hemodialysis as a treatment method. This incident underscores the need for awareness of the potential risks associated with PG and PO in different biological systems.

Another case report highlighted renal failure and polyneuropathy following ingestion of a product containing DPG (Nelsen et al. 2009).

These results show that complex biological factors can lead to a wide diversity of outcomes

### Biological Factors

The levels of propylene oxide in exhaled breath and other tissues can be influenced by various factors. These include occupational exposure, environmental conditions (such as proximity to industrial sites and air quality), lifestyle factors (such as smoking), individual metabolic rates, pre-existing health conditions, liver function, circulation, pregnancy and menstrual cycle status, age, sex, genetic makeup, microbiome composition, and nutrition.

One source of Propylene Oxide in the human body is inhalation of Propylene. PE is an odorless gas that is ubiquitous in the environment, arising from both natural and anthropogenic sources (Morgott 2018). However, the body’s conversion of propylene to propylene oxide appears to reach saturation at high exposure levels (Pottenger 2007). Individual responses to propylene exposure depend on several biological factors. Species plays a key role. Rats and mice metabolize propylene at a much higher rate than humans, leading to higher internal doses and greater health risks at the same external exposure (Filser et al. 2008). Within the same species, sex may also influence propylene metabolism and effects. Females tend to have higher respiratory rates, which can increase the internal dose (Morgott 2018).

Genetics likely influence how individuals metabolize and respond to propylene. Enzymes involved in propylene metabolism and its byproducts exhibit polymorphism, with certain variants being more active or efficient than others. The conversion of propylene oxide from propylene was confirmed in vitro using cytochrome P450, a crucial group of enzymes primarily found in the liver, playing a key role in detoxification and metabolizing various foreign compounds. This process was also demonstrated using liver microsomes and an NADPH-generating system (Groves et al., 1986; Filser et al., 2008). The most likely cytochrome from the CYP family involved in this process is CYP2E1 (rabbit liver microsomal cytochrome P450LM2), as utilized by Groves et al. (1986) and Heimbrook et al. [20]. Certain alleles in CYP2E1 gene are linked to several diseases, including hepatic cirrhosis and certain cancers, through increased biotransformation of procarcinogens. Factors that impact gene expression for some genetic variations include ethanol consumption, exposure to isoniazid and various chemicals or conditions like obesity.

Some mutations reduce the epoxidation capacity for specific substrates, affecting detoxification pathways. Genetic variations in CYP2E1 can alter the structure and function of CYP enzymes, leading to either decreased (or diminished) epoxidation resulting in lower production of epoxide metabolites, potentially compromising the detoxification of certain chemicals or increased epoxidation leading to higher levels of undesirable epoxide metabolites, potentially increasing the risk of DNA damage and cancer. Genetics of other cytochromes has been linked to hypertension, coronary artery disease, ichthyosis, corneoretinal dystrophy, increased bone density, adrenal insufficiency or hyperplasia, cholestasis, spastic paraplegia, corticosterone methyl oxidase deficiency, aromatase deficiency, vitamin D dysregulation, hypercalcemia and many others (Nebert et al. 2013)

PO is metabolized in living organisms primarily through enzymatic reactions facilitated by the liver. The metabolism of PO involves its conversion into a less reactive and less toxic diol form, 1,2-propanediol, which is then further metabolized to lactic and pyruvic acids or excreted as is. The main enzyme responsible for this conversion is epoxide hydrolase (EH), also known as microsomal epoxide hydrolase (mEH). This enzyme is found in the endoplasmic reticulum of cells and plays a crucial role in detoxifying various epoxides. Epoxide hydrolases catalyze the hydrolysis of epoxides to their corresponding diols, and it can also convert PO to propylene glycol. Several single nucleotide polymorphisms (SNPs) in the mEH gene have been identified, with some impacting enzyme activity. For example, the mEH Tyr61Ser polymorphism (rs2285097) has been associated with altered mEH activity and increased susceptibility to certain cancers.

In some cases, cytochrome P450 enzymes can also metabolize epoxides. Several biological variables, including genetic polymorphisms and the human microbiome, could modulate individual susceptibility to PO.

Individuals with a null genotype for GSTT1 may experience slight alterations in propylene oxide metabolism because GSTT1 indirectly influences the activity of other enzymes like mEH. Glutathione transferase theta (GSTT1–1) exhibits selectivity in metabolizing PO over other epoxides, suggesting a specific enzymatic preference that could impact detoxification capabilities (Theis et al., 1999). Variants in GSTT1 and GSTM1 can affect PO detoxification, with null genotypes potentially increasing susceptibility to its toxic effects (Ogawa et al., 2006). Faller et al., 2001) investigated the inactivation kinetics of propylene oxide by GST and EH enzymes in various tissues of mice, rats, and humans. Their findings revealed tissue and species-specific variations in GST and EH activities, with notable differences in metabolic rates and enzyme efficiencies across liver, lung, and nasal mucosa. The highest activities were observed in the nasal mucosa of rats for both GST and EH enzymes, significantly exceeding the levels in rat liver. Additionally, among liver and lung tissues, the highest GST activity was found in the lung of mice, while the highest EH activity was observed in human tissues. Investigation of metabolic inactivation of propylene oxide in different species indicated that hemoglobin adducts serve as a sensitive dosimeter for systemic exposure, but are not predictive of DNA binding in individual tissues.

The probable reaction pathways for propylene epoxidation with O2 and H2 in the presence of trimethylamine suggest that trimethylamine could participate in propylene oxide reactions. Utilizing trimethylamine as a gas-phase promoter for nanoparticulate gold catalysts enables direct one-stage production of PO with steady space–time yields comparable to those of current industrial ethylene oxide production. Trimethylamine may facilitate enzymatic reactions but it is not clear how this process would work. Trimethylamine is a substrate for flavin-containing monooxygenase 3 (FMO3), also known as dimethylaniline monooxygenase [N-oxide-forming] 3 and trimethylamine monooxygenase, which is a flavoprotein enzyme (EC 1.14.13.148) encoded by the FMO3 gene in humans. The FMO3 enzyme is part of a larger enzyme family called flavin-containing dimethylaniline monooxygenases (FMOs) breaking down nitrogen-containing compounds derived from the diet. The FMO3 enzyme is involved in processing certain drugs, including propylene glycol, suggesting that genetic variations in FMO3 could influence PO detoxification.

Individuals with certain genetic polymorphisms are at higher risk of adverse effects from the same propylene exposure. The body’s physiological state influences its handling of propylene. Factors like age, health status, pregnancy and menstruation cycle can alter respiration rate, circulation, liver function and other parameters in ways that change the internal propylene dose and effects (Morgott 2018). For example, higher respiration and circulation during exercise or fever can increase the internal dose. Propylene inhalation also depends on the particle size, with smaller particles penetrating deeper into the lungs. The site of respiratory tract exposure impacts the amount of propylene that reaches the bloodstream and internal organs (Wieslander 2001). In addition, the frequency and duration of exposure play a key role. Intermittent or short-term exposures are less likely to saturate metabolic pathways, allowing higher internal doses (Morgan 2011).

1,3-propanediol conversion to PO is theoretically possible but not proven. It’s most likely to occur via microbial fermentation. Alcohol dehydrogenase (ADH), Aldehyde dehydrogenase (ALDH), and Glycerol dehydratase (GDHt) could be involved. We that protein engineering, particularly directed evolution, experiments showed that CYPs can be transformed into efficient alkane hydroxylases after only five generations of random mutagenesis and recombination, optimizing epoxidation of specific substrates.

Exposure to environmental epoxides can have significant impacts on biological systems and disease susceptibility, though individual responses vary widely based on biological factors. Genetics and epigenetics play a key role in determining how individuals respond to epoxide exposures. An individual’s toxicokinetics, or how the body absorbs and distributes epoxides, also influences their response. Metabolism of epoxides by enzymes like epoxide hydrolase is key, and can vary based on factors like nutrition, chemical exposures, and genetics (Moody 1991). Receptor-mediated effects, where epoxides bind to cellular receptors, are another biological factor impacting individual responses. Schug (2011) notes that epoxides can interfere with the body’s endocrine system by blocking or mimicking the actions of natural hormones. This endocrine disruption can then lead to disease endpoints like reproductive disorders or metabolic disease. Developmental stage during exposure is critical in determining impacts. Exposure to epoxides during early development can alter disease susceptibility later in life (Schug 2011). Epigenetic changes caused by developmental epoxide exposure may even persist across generations (Guerrero-Bosagna 2014).

In summary, biological factors like species, sex, genetics, physiology and exposure characteristics, including duration and co-exposure to other chemicals, significantly influence how individuals metabolize propylene oxide. An individual’s toxicokinetics, genetics, receptor-mediated effects, and developmental stage all interact to influence how their biology responds when exposed to environmental epoxides. Seasonal fluctuations in PO exposures have been also noted, possibly because sporce VOCs are more easily to be evaporated in higher temperature and inhalation is their primary human exposure route (Li et al. 2023). These complex biological factors lead to a diversity of outcomes, from adaptation to disease. A deeper understanding of these factors will enable more accurate assessment and prediction of the impacts of epoxide exposures on human health, as well as the use of measured values as biomarkers of other processes in the body.

### Microbial involvement

Microbial involvement in the metabolism and detoxification of propylene oxide has been observed in various studies. Certain microbes, such as Pseudomonas aeruginosa, possess unique metabolic pathways capable of metabolizing PO (Huybregtse and Vanderlinden, 1964).

Methanotrophic bacteria, such as Methylococcus capsulatus (Bath), have been shown to oxidize gaseous alkenes, including propylene, to the corresponding epoxides utilizing an NADH2-dependent methane monooxygenase. The ability of methanotrophic bacteria to produce propylene oxide with formate, methanol, and methane was also mentioned.

These pathways are not typically present in the human microbiome, suggesting that microbial colonization could influence the overall metabolism of PO in the body. Genetic variations in human-microbe interactions could potentially impact the efficiency of PO degradation or transformation, leading to varying levels of exposure and potential toxicity.

Escherichia coli has been shown to increase the yield of 1,2-propanediol when the methylglyoxal synthase gene (mgs) is overexpressed, indicating a potential microbial contribution to the detoxification process of PO. Flavobacterium found in human specimens can grow on grow on the diol as the sole source of carbon and could be involved in Propylene Oxide metabolism. The conversion of PO into propylene glycol by bacteria and catalysts suggests an intrinsic ability within both microbial life and chemical processes to detoxify and transform PO.

Specific bacterial strains like Xanthobacter autotrophicus Py2 and Rhodococcus rhodochrous B276 have been identified to metabolize propylene through a coenzyme M (CoM)-dependent pathway, which is not commonly found in the human microbiome. This pathway involves the transformation of propylene oxide into acetoacetate, with CoM acting as a key cofactor in the epoxide ring opening.

Propylene oxide saponification wastewater is enriched by bacterial genus Azoarcus (plant microbiome) and many bacterial viruses, indicating that there could be complex pathways and dependences connected to Propylene Oxide, its precursors and derivatives.

Microbial involvement in the metabolism and detoxification of propylene oxide has been observed in various studies. Certain microbes, such as Pseudomonas aeruginosa, possess unique metabolic pathways capable of metabolizing PO (Huybregtse and Vanderlinden, 1964).

Methanotrophic bacteria, such as Methylococcus capsulatus (Richards et al. 1994), have been shown to oxidize gaseous alkenes, including propylene, to the corresponding epoxides utilizing an NADH2-dependent methane monooxygenase. The ability of methanotrophic bacteria to produce propylene oxide with formate, methanol, and methane was also mentioned.

These pathways are not typically present in the human microbiome, suggesting that microbial colonization could influence the overall metabolism of PO in the body. Genetic variations in human-microbe interactions could potentially impact the efficiency of PO degradation or transformation, leading to varying levels of exposure and potential toxicity.

Escherichia coli has been shown to increase the yield of 1,2-propanediol when the methylglyoxal synthase gene (mgs) is overexpressed, indicating a potential microbial contribution to the detoxification process of PO. Flavobacterium found in human specimens can grow on diols as the sole source of carbon and could be involved in propylene oxide metabolism. The conversion of PO into propylene glycol by bacteria and catalysts suggests an intrinsic ability within both microbial life and chemical processes to detoxify and transform PO.

Specific bacterial strains like Xanthobacter autotrophicus Py2 and Rhodococcus rhodochrous B276 have been identified to metabolize propylene through a coenzyme M (CoM)-dependent pathway, which is not commonly found in the human microbiome. This pathway involves the transformation of propylene oxide into acetoacetate, with CoM acting as a key cofactor in the epoxide ring opening.

Populations in Salmonella and other Gram-negative bacteria significantly decrease in response to PO exposure, especially as the relative humidity of the air within the chamber containing the propylene oxide vapor are increased (Himmelfarb et al. 1962). While Salmonella lacks cytochrome P450 enzymes typically involved in epoxide metabolism, it possesses glutathione S-transferases (GSTs) and epoxide hydrolases (EH). PO’s toxicity, however, could be direct - disrupting membranes and damaging DNA and proteins before its enzymes can intervene.

Propylene oxide saponification wastewater is enriched by the bacterial genus Azoarcus (plant microbiome) and many bacterial viruses, indicating that there could be complex pathways and dependencies connected to propylene oxide, its precursors, and derivatives (Fan et al. 2023).

## Discussion

### Principal Findings

The systematic review of propylene oxide research highlights a shift in focus over recent years, with increasing recognition of its public health significance versus occupational exposure. Inhalation remains the primary route of occupational exposure to PO, notably in industries utilizing it for polyurethane polyols, propylene glycol production, or as a fumigant. The only publicly available report on levels of PO in exhaled human breath (Filser et al, 2008: 0.083-0.3 ppb), holds pivotal significance for analyzing data in the general population This significance is further underscored by our own study, which identified higher levels of PO in a subset of environmentally sensitive individuals, suggesting individual metabolic variations that impact PO exposure and toxicity [5,16].

Our findings stress the importance of considering individual variability in response to PO exposure, influenced by both genetic and environmental factors. Various publications emphasize the intricate interplay between environmental exposures, genetic makeup, and individual susceptibility regarding biological factors impacting PO levels. Propylene Oxide acts as a direct-acting alkylating agent, exerting genotoxic effects on DNA. Metabolism predominantly occurs through glutathione conjugation and epoxide hydrolase-mediated hydration, with interspecies differences highlighted, necessitating caution when extrapolating findings from animal models to human risk assessment.

Additionally, the human microbiome’s influence on PO metabolism further underscores the complexity of individual responses. Environmental factors, including exposure to other chemicals and seasonal variations, also modulate PO’s impact, necessitating comprehensive assessments in exposure evaluations.

Understanding the intricate interplay of biological and environmental factors in PO metabolism and toxicity is crucial for accurate risk assessment and intervention development. Future research should focus on mechanisms driving individual variability in response to PO and exploring gene-environment interactions.

### Advancing through AI

The utilization of AI tools such as Elicit, ChatGPT, Gemini, and Perplexity has shown promise in identifying pertinent literature, highlighting the increasing integration of technology in systematic reviews. These AI tools keep improving in identifying the best references and multi-agent flows hold great promise. However, challenges persist, particularly in capturing nuanced scientific data and ensuring consistency across platforms. Although AI excels in refining language and enhancing readability, and is becoming increasingly adept at processing larger texts, it may still miss critical information due to its reliance on explicitly provided data. Humans, on the other hand, can leverage implicit knowledge and experience to identify and prioritize key details that AI might overlook. This gap suggests that AI needs to enhance its abilities to critically process new information by comparing old and new data and discerning between relevant and less relevant content.

This dynamic nature of scientific literature presents a hurdle for knowledge engineering in systematic reviews, as evidenced by the struggles of sustaining “live reviews” or dynamic indexing, as observed in the case of MEDLINE database searches for PubChem IDs (unfortunately, not updated since 2012).

To address these limitations, future AI development should focus on improving the ability to critically analyze and compare new information with existing knowledge. Enhancing AI’s capability to understand and process nuanced information will be vital. This might involve creating specialized AI models that can dynamically update reviews and integrate multi-agent flows to provide more comprehensive insights.

### Limitations

The accuracy and comprehensiveness of our findings are contingent upon the quality and completeness of the original publications. Variability among the included reviews in terms of focus, scope, methodology, and quality may limit the applicability of our results to a broader context.

This systematic review was conducted by one human author in collaboration with several AI agents, potentially introducing subjective interpretations. While AI tools can aid in data extraction and analysis, human oversight remains essential to ensure the reliability and validity of the review process.

Analyzing propylene oxide exposures across different studies presents complexities due to variability in concentration, exposure duration, species, and observed outcomes. The included studies demonstrate a wide range of PO concentrations and exposure periods, involving various organisms, complicating direct comparisons and quantitative synthesis of results.

## Conclusions

This systematic review sheds light on the evolving landscape of propylene oxide research, emphasizing its growing public health significance beyond occupational exposure. Key findings highlight the routes of PO exposure across various settings, along with identified enzymes and microbial associations. The importance of individual variability in response to PO exposure, influenced by factors like sex, dietary preferences, genetics, and physiology, has been underscored. This review also points to the intricate interplay between environmental exposures, genetic makeup, and individual susceptibility in determining PO levels in the body.

The increasing role of AI in advancing systematic reviews is undeniable, and the future lies in optimizing these tools to address their current limitations. Developing specialized AI systems for dynamic reviews can keep research updated in real-time, potentially through specialized models designed for systematic reviews. Moreover, integrating multi-agent AI flows can enhance the accuracy and depth of literature reviews, ensuring comprehensive and precise identification of relevant studies.

Future research should continue to explore the interplay between genetic and environmental factors influencing PO metabolism. Understanding individual susceptibility to PO exposure, especially among environmentally sensitive individuals, will be crucial for developing targeted interventions and policies to mitigate health risks. As AI continues to evolve, its integration with human expertise will be essential to advance our understanding of environmental exposures and its implications for public health.

## Acknowledgements

The author thanks ChatGPT and Bard (now Gemini) for their collaborative efforts during the review process. AI tools played a crucial role as independent labelers and validators, ensuring both factual accuracy and grammatical correctness. Following this, the human author meticulously re-evaluated all facts and papers identified as relevant by the AI tools.

## Data Availability Statement

Data is available as open access at OSF: https://osf.io/w7682/

## Abbreviations

2-HPMA: N-acetyl-S-(2-hydroxypropyl)cysteine, mercapturic acids derived from propylene oxide
ADH: Alcohol dehydrogenase
AEGL: Acute Exposure Guideline Levels developed by EPA
AI: Artificial Intelligence
ALDH: Aldehyde dehydrogenase
CYPs: Cytochromes P450, involved in the metabolism of xenobiotics in the body
CYP2E1: Cytochrome P450 2E1, a member of the cytochrome P450 mixed-function oxidase system
EH: epoxide hydrolase, also known as mEH
EPA: the Environmental Protection Agency
FMO: flavin-containing monooxygenase enzyme family
GST: Glutathione S-transferase family
IARC: International Agency for Research on Cancer
LC50: median lethal concentration in air that can kill 50% of a test population
LD50: median lethal dose that can kill 50% of a test population
LOA: Level of Distinct Odor Awareness
mEH: microsomal epoxide hydrolase
MeSH: Medical Subject Headings, a controlled and hierarchically-organized vocabulary produced by NLM
NLM: the National Library of Medicine
NPSH: nonprotein sulfhydryl (SH-groups)
OSF: Open Science Framework, a platform for data sharing and collaboration
PPB: parts per billion
PPBV: parts per billion by volume, specifically refers to the concentration of a substance in a gaseous mixture
PPM: parts per million
PPMV: parts per million by volume
PRISMA: Preferred Reporting Items for Systematic Reviews and Meta-Analyses
PE: Propylene
PG: Propylene Glycol
PO: Propylene Oxide
RNM: respiratory nasal mucosa
RQ: Research Question
SCE: sister chromatid exchanges
SMR: standardized mortality ratio;
SSB: DNA single-strand breaks
TWA: time weighted average
URT: upper respiratory tract
US EPA: US Environmental Protection Agency
VOSviewer: “Visualization of Similarities Viewer” - a software tool developed for constructing and visualizing scientific landscapes

